# An Analysis of Transgender Crowdfunding in the Context of State-Led Anti-Transgender Legislation

**DOI:** 10.1101/2024.09.19.24313982

**Authors:** Meera Nagpal, Julia Wang, Anjali Mahapatra, Neha Chintapally, Christi Butler

**Affiliations:** USF Morsani College of Medicine; University of California San Francisco

## Abstract

This study investigates the impact of anti-transgender legislation on the use of crowdfunding platforms by transgender and gender diverse (TGD) individuals in the United States. Using data from 349 GoFundMe campaigns created between March and June 2022, we analyzed the reasons for seeking funding and their potential correlation with state-level anti-transgender policies. Our findings reveal that 61% of campaigns originated from states with proposed or enacted anti-transgender legislation, with 19.5% coming from states posing significant barriers to healthcare services for TGD individuals. The most common reasons for crowdfunding included housing (44.7%), living expenses (35.5%), and gender-affirming surgeries (28.1%). Notably, campaigns from anti-transgender states showed a significant correlation with explicitly mentioning hostile state environments (Φ = 0.2843, p < 0.0001) and seeking relocation (Φ = 0.2040, p = 0.0001). These results highlight the profound impact of discriminatory legislation on TGD individuals, forcing many to seek alternative financial support for basic needs and essential medical care. This research underscores the urgent need for more inclusive healthcare policies and support systems for the TGD community.

## Introduction

Access to health care has historically been a challenge for some of the most vulnerable populations in the United States. This has included transgender and gender diverse (TGD) individuals who have faced years of mistreatment and isolation from the healthcare system^1^. In more recent years it seems that care for TGD individuals has become more highly politicized with anti-transgender legislation on the rise^2-6^. In 2018, 30 anti-transgender state bills were proposed, compared to 2022, when the number of proposed bills jumped to 174, and in 2023, 615 anti-transgender bills were introduced (Figure 1)^9,10^. These legislative measures range from participation restrictions on trans-identifying girls and women in female sports, prohibiting TGD youth from accessing bathrooms and locker rooms corresponding to their gender identity, and imposing limitations on gender-affirming medical treatment^3,9^. These limitations have included prohibiting provisions of gender-affirming care and even criminalizing physicians who provide such care^4,7-10^. The impact of such laws has only resulted in creating significant barriers to healthcare services including access to hormone therapy, gender-affirming surgeries, and mental health support^3-5,7,8^.

**Figure 1.**
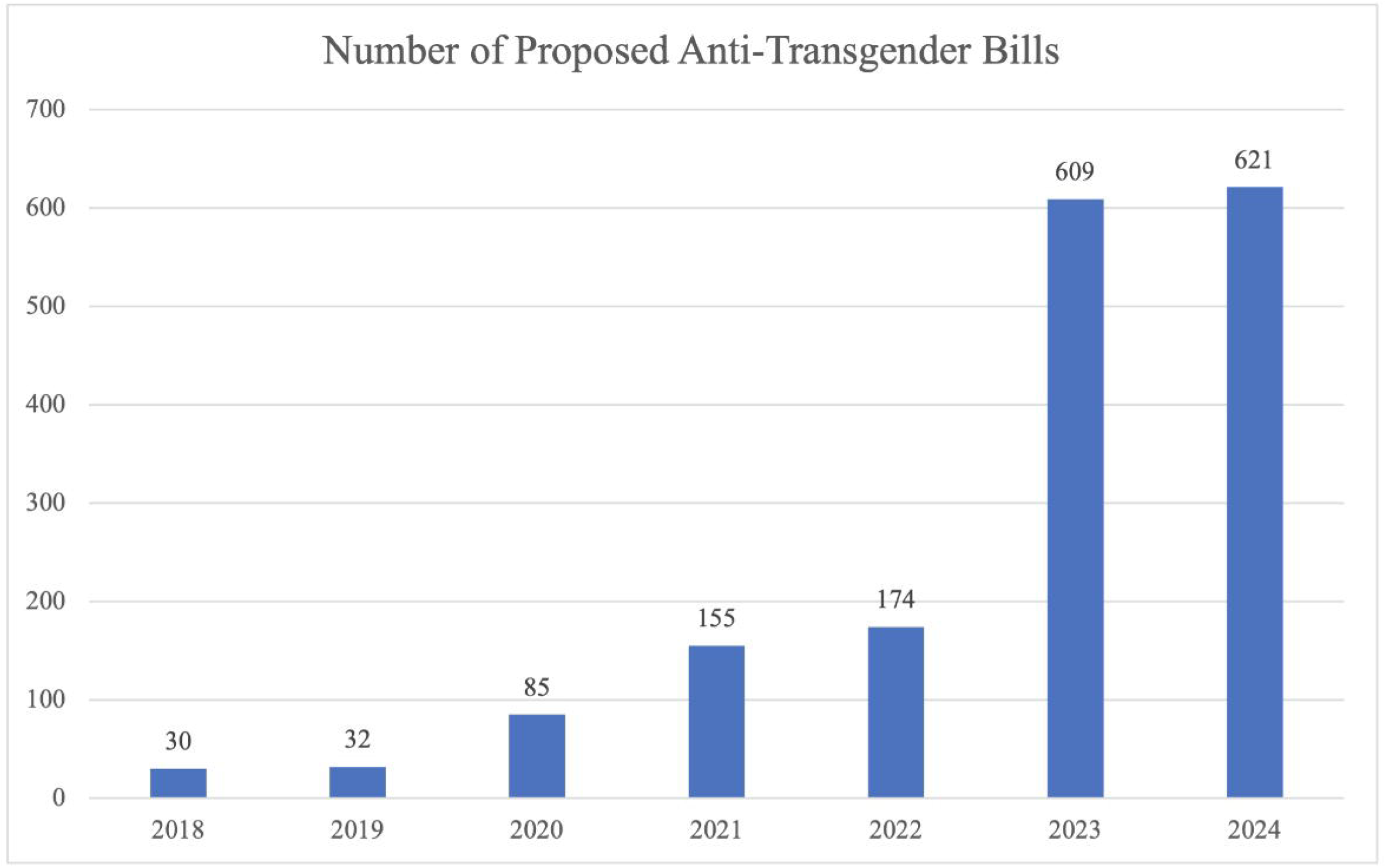
Number of Proposed Anti-Transgender Bills

In response to these barriers, individuals may seek alternative resources and measures to meet their healthcare needs. Unfortunately, these alternatives may be unsanctioned and potentially hazardous, posing significant risks to their health and safety^17,18^. The lack of regulated and safe options not only endangers their well-being but can also result in additional, often substantial, financial costs^19^.

Crowdsourcing platforms have emerged as a popular avenue for individuals to raise funds for various needs, including costs related to health care^20^. With the escalation of anti-transgender legislation in the United States, it is likely that TGD individuals from states with such legislation may increasingly rely on web-based crowdfunding to finance healthcare expenses and gender related issues^21,22^.

This study seeks to investigate whether the implementation of state anti-transgender laws (either proposed or passed) had any influence on the use of and/or reason for seeking crowdfunding platforms among trans-identifying individuals. The goal of this research is to shed light on the financial struggles faced by TGD individuals residing in these anti-inclusive legislative environments and provide insights into the potential impact of anti-transgender legislation on crowdfunding trends within TGD communities.

## Methods

Using the crowdsourcing platform, GoFundMe.com (GFM), we set out to understand why TGD individuals sought funding and if there was any connection to living in a state that supported anti-transgender legislations. The selection of GFM was based on its status as the largest online crowdfunding platform and its popularity within TGD communities^23,24^. As the data accessed was publicly available, it did not fall under the category of human subject data, as defined by institutional review boards and was exempt from IRB approval. We present the data in aggregated form to safeguard the privacy of fundraisers.

In June 2022, GFM was queried for active campaigns containing the term “trans”, which yielded 1000 search results^25^. This was then narrowed to include active campaigns created for individual beneficiaries identifying as transgender seeking financial support. Campaigns that occurred outside of the United States, from organizations, or not related to transgender individuals were excluded. This resulted in a final count of 349 relevant campaigns posted between March 2022 and June 2022. A dataset including author demographics and narrative details was compiled.

Through the process of inductive coding, researchers conducted a first pass through the data to explore themes present in the campaign title and narrative^26^. Codes were assigned to identified themes including medical treatment, personal finances, abuse, discrimination, and mental health. Codes were selected based on discussion and agreement amongst all authors. Codes were subsequently reviewed by at least 2 authors to ensure reproducibility and consensus in data extraction. State of residence, gender identity, and race were obtained as self-described on the campaign page.

The Trans Legislation Tracker, an independent research organization, as well as the American Civil Liberties Union (ACLU), were utilized as the primary data sources for identifying and categorizing anti-transgender legislation across the United States. These resources systematically monitor and document bills that impact TGD individuals, providing a comprehensive database of proposed and enacted laws^5,6^. For the purposes of this study, researchers used data from the Trans Legislation Tracker to establish a quantitative measure of anti-transgender legislative activity by state for the year of 2022^6^. Additionally, anti-transgender legislation from 2021 was collected from ACLU^5^. States were classified as “anti-transgender” if they had enacted bills that restrict TGD rights or access to services for the purposes of statistical correlation analysis.

To analyze the relationship between anti-transgender state status and thematic codes from crowdfunding campaigns, we used Phi coefficients (Φ) to measure the strength of association between these binary variables. Statistical significance was assessed using chi-square tests of independence, with Fisher’s exact tests employed for cases with low expected frequencies. We set the significance level at p < 0.05. All analyses were conducted using Python 3.9^27^, with pandas library (version 1.3.4)^28^ for data manipulation and organization, numpy (version 1.22.4)^29^ for numerical operations, and scipy.stats (version 1.7.1)^30^ for statistical computations.

## Results

After review, it was found that many campaigns commonly requested funding for medical expenses, pre-surgical requirements such as hair removal, travel to obtain medical care, and relocation due to safety (Table 1). Specifically, results indicated that the majority of campaigns were used to fund housing, (44.7%, n=156), living expenses (35.5%, n=124), gender-affirming surgery (28.1%, n=98), hormone therapy (17.8%, n=62), and general healthcare costs outside of gender-affirming care (13.5%, n=47).

**Table 1:** Funding Needs in GoFundMe Campaigns.

| Funding Need | n | % |
| --- | --- | --- |
| Housing | 156 | 44.7% |
| Living expenses | 124 | 35.5% |
| Surgery | 98 | 28.1% |
| Hormones | 62 | 17.8% |
| Non-gender-affirming medical care | 47 | 13.5% |
| Transportation | 43 | 12.3% |
| Gender-affirming healthcare, unspecified | 36 | 10.3% |
| Hair removal | 33 | 9.5% |
| Relocation, out-of-state | 28 | 8.0% |
| Debt | 19 | 5.4% |
| Therapy | 13 | 3.7% |
| Legal fees for name change | 10 | 2.9% |

Analysis of campaign narratives revealed several prevalent themes, including mental health concerns (23.2%, n=81), experiences of abuse (16.3%, n=57), gender dysphoria (17.2%, n=60), and exposure to hostile environments (35.2%, n=123) (Table 2).

**Table 2:** Themes in GoFundMe Campaigns.

| Theme | n | % |
| --- | --- | --- |
| Mental illness | 81 | 23.2% |
| Abuse | 57 | 16.3% |
| Dysphoria | 60 | 17.2% |
| Hostile environment | 123 | 35.2% |
| Houseless/homeless | 42 | 12.0% |
| Insurance: gender-affirming healthcare | 41 | 11.7% |
| Disabled | 44 | 12.6% |
| Hostile state | 27 | 7.7% |
| Neurodivergence | 21 | 6.0% |
| Chronic illness | 12 | 3.4% |

Results from GFM campaigns reveal that 61% (n=213) of campaigns came from states that have passed or proposed anti-transgender legislation. This was correlated with data collected from 2022 and 2021, which revealed 33 states that initiated anti-transgender legislation, with 13 states having passed bills or adopted policies, and 1 directive from the Governer of Texas declaring gender-affirming care to be considered illegal or child abuse (Table 3)^5,6^. 19.5% of GoFundMe campaigns originate from states with specific policies that pose significant barriers to healthcare services for TGD individuals.

**Table 3:** States and Anti-Transgender Legislation Status from 2021-2022.

| Passed | Proposed |
| --- | --- |
| Alabama | Alaska |
| Arizona | Delaware |
| Florida | Hawaii |
| Georgia | Idaho |
| Indiana | Illinois |
| Iowa | Kansas |
| Kentucky | Maryland |
| Louisiana | Minnesota |
| Oklahoma | Mississippi |
| South Carolina | Missouri |
| South Dakota | New Hampshire |
| Tennessee | New Jersey |
| Texas | Ohio |
| Utah | Pennsylvania |
|  | Rhode Island |
|  | Virginia |
|  | Washington |
|  | West Virginia |
|  | Wisconsin |
|  | Wyoming |

7.7% of campaigns (n=27) specifically ascribed living in a hostile state as a significant reason for requesting funding. This was cross referenced with anti-transgender legislation as documented on Table 3. It was found that all campaigns in this population were from authors based in states that have passed anti-transgender legislation. These include Alabama, Florida, Georgia, Texas, South Carolina, and West Virginia, and Arkansas (Table 4)^5,6^.

**Table 4:** GoFundMe Campaign Recipient State of Origin.

| State | n | % | State | n | % |
| --- | --- | --- | --- | --- | --- |
| Alabama | 8 | 2.3% | Montana | 0 | 0.0% |
| Alaska | 1 | 0.3% | Nebraska | 1 | 0.3% |
| Arizona | 8 | 2.3% | Nevada | 3 | 0.9% |
| Arkansas | 3 | 0.9% | New Hampshire | 1 | 0.3% |
| California | 38 | 10.9% | New Jersey | 10 | 2.9% |
| Colorado | 8 | 2.3% | New Mexico | 2 | 0.6% |
| Connecticut | 4 | 1.1% | New York | 25 | 7.2% |
| Delaware | 1 | 0.3% | North Carolina | 7 | 2.0% |
| Florida | 26 | 7.4% | North Dakota | 0 | 0.0% |
| Georgia | 17 | 4.9% | Ohio | 10 | 2.9% |
| Hawaii | 2 | 0.6% | Oklahoma | 4 | 1.1% |
| Idaho | 0 | 0.0% | Oregon | 13 | 3.7% |
| Illinois | 18 | 5.2% | Pennsylvania | 15 | 4.3% |
| Indiana | 3 | 0.9% | Rhode Island | 2 | 0.6% |
| Iowa | 2 | 0.6% | South Carolina | 2 | 0.6% |
| Kansas | 4 | 1.1% | South Dakota | 0 | 0.0% |
| Kentucky | 2 | 0.6% | Tennessee | 4 | 1.1% |
| Louisiana | 1 | 0.3% | Texas | 23 | 6.6% |
| Maine | 4 | 1.1% | Utah | 4 | 1.1% |
| Maryland | 8 | 2.3% | Vermont | 1 | 0.3% |
| Massachusetts | 7 | 2.0% | Virginia | 7 | 2.0% |
| Michigan | 9 | 2.6% | Washington | 14 | 4.0% |
| Minnesota | 6 | 1.7% | West Virginia | 1 | 0.3% |
| Mississippi | 0 | 0.0% | Wisconsin | 4 | 1.1% |
| Missouri | 7 | 2.0% | Wyoming | 0 | 0.0% |
| Washington, DC | 5 | 1.4% | Unknown | 4 | 1.1% |

11.5% of campaigns (n=41) cited lack of insurance coverage of gender-affirming healthcare for their reasoning seeking funding, and states in this population include Missouri, Minnesota, Arizona, Virginia, Georgia, and Texas, all of which also have current or actively proposed legislation that is viewed as anti-transgender^5,6^.

The Phi coefficient analysis revealed varying levels of correlation between GoFundMe campaign origins in anti-transgender states and the specific codes representing different needs (Table 5). Of particular note is the statistically significant correlation between campaign origin in anti-transgender states and the “Hostile State” code (Φ = 0.2843, p = 0.0000). Additionally, the “Relocation Out of State” theme showed significant correlations with anti-transgender state status (Φ = 0.2040, p = 0.0001).

**Table 5:** Correlations between Codes and Anti-Transgender State Status.

| Code | Phi Coefficient | p-value |
| --- | --- | --- |
| Housing | 0.0166 | 0.7565 |
| Living Expenses | 0.0587 | 0.2732 |
| Surgery | 0.0214 | 0.6896 |
| Hormones | 0.0083 | 0.8772 |
| Non-gender-affirming medical care | 0.0381 | 0.4772 |
| Transportation | 0.0886 | 0.0981 |
| Gender-affirming healthcare, unspecified | 0.0707 | 0.1863 |
| Hair removal | 0.0344 | 0.5210 |
| **Relocation, out-of-state** | **0.2040** | **0.0001\*** |
| Debt | 0.0459 | 0.3915 |
| Therapy | 0.0163 | 0.7607 |
| Legal fees for name change | 0.0000 | 1.0000 |
| Mental illness | 0.0196 | 0.7138 |
| Abuse | 0.0508 | 0.3422 |
| Dysphoria | 0.0147 | 0.7829 |
| Hostile environment | 0.0103 | 0.8479 |
| Houseless/homeless | 0.0645 | 0.2282 |
| Insurance: gender-affirming healthcare | 0.0400 | 0.4554 |
| Disabled | 0.0934 | 0.0809 |
| **Hostile state** | **0.2843** | **0.0000\*** |
| Neurodivergence | 0.0277 | 0.6043 |
| Chronic illness | 0.0402 | 0.4525 |

\*Statistically significant ( $p < 0.05$ )

## Discussion

The role of crowdfunding within TGD communities remains largely underexplored^21,22,31-33^. This study looked at associations between the existence of anti-transgender legislation and the use of crowdfunding specifically for themes that may be impacted by anti-transgender legislation such as access to medical care and safety.

Crowdfunding, through platforms like GoFundMe and social media, has become an efficient and popular way to raise funds for a variety of reasons including medical costs for gender affirming surgery. A recent study analyzed GoFundMe campaigns for gender affirming surgery in the United States from 2012-2016 and discovered that the majority of campaigns were used to fund chest surgeries for young, white, binary-identified trans men. Few campaigns reached their goal, and those that did tended to have more shares on social media and older recipients with a larger social network^21^. They concluded that increasing reliance on crowdfunding platforms as a means to facilitate access for gender affirming healthcare has inherent limitations, such as loss of privacy and disparities in accessibility especially by TGD individuals^21,22,31^. Our study furthers these findings and supports that trans-exclusionary policies and practices force individuals to turn to other resources to fund or even provide their medical care.

Our analysis of crowdfunding campaigns reveals the significant impact of anti-transgender legislation on the TGD community. Anti-transgender legislation has driven increased reliance on crowdfunding platforms for gender-affirming healthcare, with 28.4% of campaigns seeking funds for surgeries and 18.1% for hormone therapy. A striking 61% of campaigns come from states with anti-transgender legislation proposed or enacted. A significant portion (19.5%) of GoFundMe campaigns originate from states with policies that create substantial barriers to healthcare services for TGD individuals. 64.2% of relocation-themed campaigns explicitly cited hostile legislative environments as the primary motivation for seeking funds to move to safer states. 11.5% of all campaigns cited lack of insurance coverage for gender-affirming healthcare as a reason for seeking funding, potentially exacerbated by discriminatory laws in certain states. The high prevalence of crowdfunding campaigns for housing (44.7%), living expenses (35.5%), and healthcare underscores how anti-transgender legislation may contribute to financial insecurity and force individuals to seek alternative means of support, basic necessities, and safer living environments.

Our analysis reveals subtle but noteworthy patterns in the relationship between anti-transgender state policies and various challenges faced by TGD individuals seeking financial support through crowdfunding campaigns. While most correlations were weak and not statistically significant, two key findings emerged: First, the “Hostile State” code showed a statistically significant positive correlation with anti-transgender state status (Φ = 0.2843, p = 0.0000). Second, the “Relocation Out of State” theme also demonstrated a significant correlation with anti-transgender state status (Φ = 0.2040, p = 0.0001). This suggests that campaigns from anti-transgender states are more likely to explicitly mention hostile state environments. Although the correlation is modest, it underscores the tangible impact of state policies on the lived experiences of TGD individuals. This finding is crucial for understanding the broader implications of anti-transgender policies and the ways in which they directly influence the lives of TGD individuals, compelling them to seek external support to navigate or escape these environments^22,34-36^.

Interestingly, factors such as housing needs, medical procedures, and healthcare access did not show significant correlations with anti-transgender state status. This could indicate that these challenges are pervasive across all states, regardless of their stance on TGD rights. Alternatively, it might suggest that the impact of state policies on these specific needs is more nuanced or indirect than initially hypothesized. The lack of strong correlations for most codes does not necessarily imply the absence of state-level effects. Instead, it highlights the complex nature of challenges faced by TGD individuals, which may transcend simple state policy categorizations. Factors like personal circumstances, local community support, and individual resilience likely play substantial roles alongside state-level policies^34,37-40^.

Anti-transgender legislation creates fear-mongering and a hostile environment for TGD individuals^2^. Studies show these laws correlate with negative mental health indicators, exacerbate health disparities, and impede transgender people’s ability to live authentically^34,36,47^. For example, internet search data analysis reveals spikes in depression and suicide-related queries following anti-transgender bills^57^, while public health research highlights increased stress and anxiety among transgender youth^58^. Conversely, most evidence suggests gender-affirming hormone therapy (GHT) is associated with better mental health outcomes, especially when initiated during adolescence^35,59^. The 2022 US Transgender Study, for example, found that 98% of respondents undergoing hormone treatment reported increased life satisfaction^34^. Anti-transgender laws often cite the Rapid Onset Gender Dysphoria (ROGD) hypothesis to justify their legislative efforts^11,12^. The ROGD hypothesis posits that there is a sudden and rapid onset of gender dysphoria, particularly among teenagers, often attributed to social contagion or peer influence rather than genuine gender identity issues^13^. Advocates of these laws argue that young people, particularly minors, should not be allowed to undergo gender-affirming medical treatments or procedures because their gender dysphoria might be a temporary phase influenced by social factors rather than a genuine identity^11,13-15^. However, there has yet to be evidence to support this hypothesis. In fact, recent research challenges the assumptions of the ROGD hypothesis by showing that a significant proportion of individuals do not realize their TGD identities until adolescence or later in life^16^. This contradicts the idea of rapid onset, suggesting instead that TGD identities can develop over time and are not solely influenced by social factors or peer pressure^11,16^.

There is a concern that the seemingly expanding anti-trans rhetoric in the U.S. will increase the health disparities gap for TGD individuals seeking medical care. There is already evidence that the hostile legislative environment in certain states has limited insurance coverage for gender affirming treatments, and the fear is this will only worsen^44,47,48^. The U.S. healthcare system is one of the most expensive in the world making up roughly twenty percent of the economy^41^. Medical bills are the number one reason for bankruptcy in America, and national healthcare costs are expected to be $6.2 trillion by 2028^41^. In addition to the cost of transitional treatments such as hormone therapy, voice coaching, hair implants, prosthetics, chest binders, and more, gender affirming medical procedures can impose significant financial burdens^45^. Genital surgeries can cost up to $26,000 while chest surgeries can reach as high as $21,000^42^. Out-of-pocket costs of hormone therapy can range from $72 to $3,792^67^. Insurance may not cover these costs as they can be viewed as medically unnecessary, a narrative supported by anti-trans legislation, leading to a large financial burden on the TGD individuals that need care^43,44^. The overall financial burden of healthcare, combined with the specific requirements for some gender-affirming surgeries, can limit access to care^1,46^. The prohibitive costs associated with these essential medical procedures can be overwhelming for many, potentially leaving them without viable options for necessary care^49-52^.

Moreover, denying access to such fundamental healthcare services exacerbates feelings of ostracism and discrimination within this population. Being unable to obtain critical medical care can profoundly impact an individual’s sense of dignity and inclusion, further marginalizing an already vulnerable group^37,51^. As a result, TGD individuals may be forced to seek unsanctioned or “DIY” treatment options to fund their healthcare needs, which carry significant health risks^21,22,32,53^. These may include obtaining hormones through unregulated sources, self-administering without proper medical supervision, or seeking surgical procedures from unqualified practitioners. Such practices can lead to severe complications, including improper dosing, infections, organ damage, and potentially life-threatening side effects^68,69^. Furthermore, the use of these unregulated treatments often precludes necessary medical monitoring, potentially exacerbating underlying health conditions or causing unforeseen medical issues^69^. Our study demonstrated, 28.4% (n=99) of crowdfunding campaigns asked for assistance funding gender-affirming surgery, and 18.1% (n=63) requested donations for hormone therapy. 11.5% (n=41) of campaigns specifically cited lack of insurance coverage for gender-affirming care. Laws restricting access to gender-affirming care may have particularly severe consequences for those with chronic illnesses or mental health conditions who rely on consistent and comprehensive healthcare^63^. While 74.5% of campaigns did not specify their race, those who did were people of color represented by 25.5% (Table 6). It is reasonable to infer that within those who did not specify race, the true number of racially diverse individuals may be higher, emphasizing the intersectionality of identities upon the experience of TGD people.

**Table 6:**
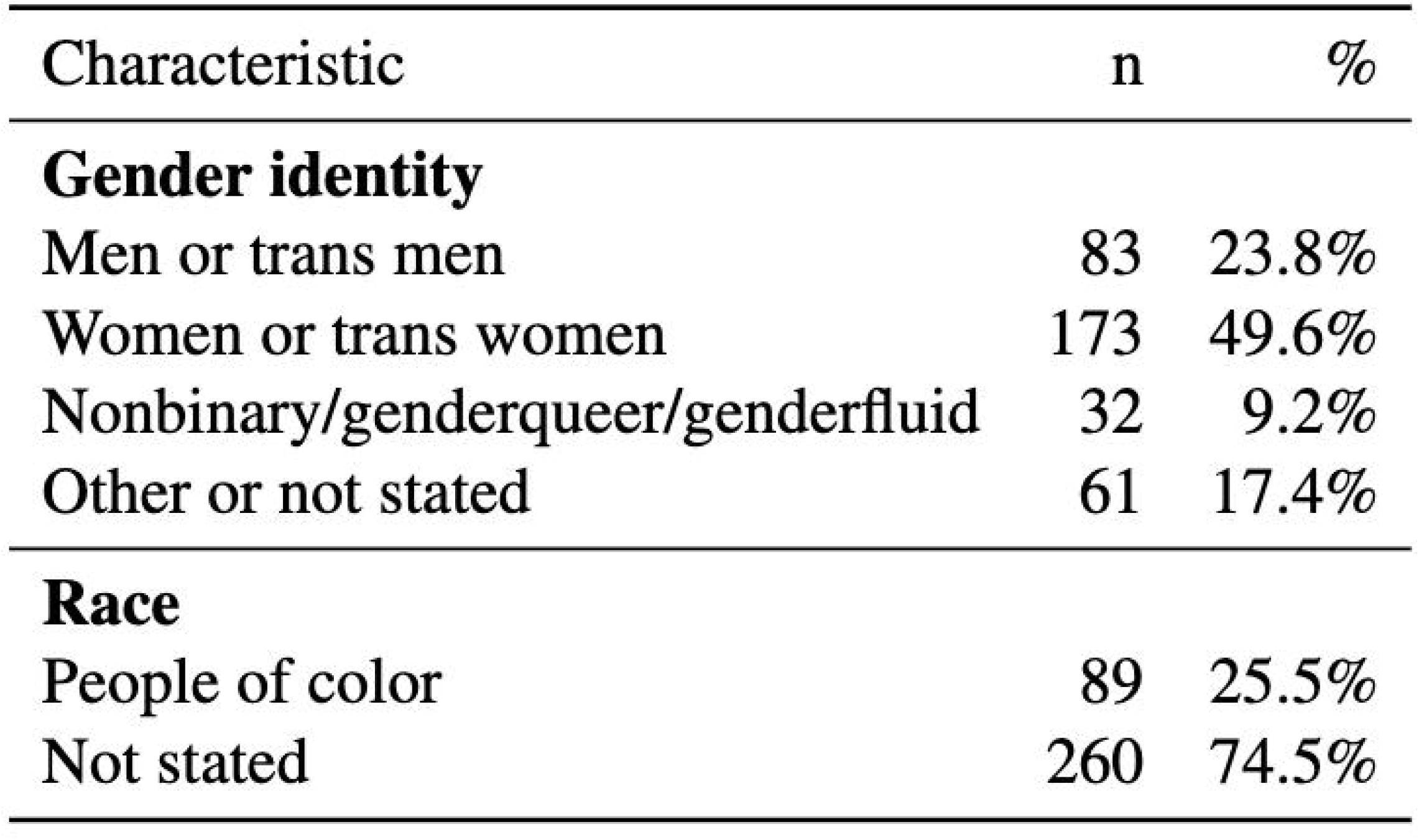
GoFundMe Campaign Recipient Characteristics.

| Characteristic | n | % |
| --- | --- | --- |
| **Gender identity** |  |  |
| Men or trans men | 83 | 23.8% |
| Women or trans women | 173 | 49.6% |
| Nonbinary/genderqueer/genderfluid | 32 | 9.2% |
| Other or not stated | 61 | 17.4% |
| **Race** |  |  |
| People of color | 89 | 25.5% |
| Not stated | 260 | 74.5% |

The impact of anti-transgender legislation extends beyond immediate financial concerns and medical care access, permeating various aspects of TGD individuals’ lives. Strikingly, 27% of campaigns cited hostile environments as a primary reason for seeking funding, indicating that challenges persist not only at the state level but also within local communities, including neighborhoods and workplaces. This pervasive hostility, potentially exacerbated by discriminatory legislation, underscores the multifaceted nature of the obstacles faced by the TGD community. The necessity for relocation due to both state-level policies and community-based hostility highlights the deep-rooted discrimination and lack of acceptance encountered in numerous facets of daily life. These hostile environments contribute to a cascade of negative outcomes, including mental health challenges, safety concerns, and diminished overall well-being, emphasizing the critical need for fostering inclusive and supportive spaces for TGD individuals across all societal levels.

There are limitations to this study given that the methods are purely observational. There can be no direct correlations drawn from this. However, the goal was to look at the trends and evaluate the possible impacts of the rise in anti-transgender legislation. There is a clear association with use of crowdfunding in the setting of anti-transgender politics, and future study should include directly evaluating the impact on the number of gender-affirming surgeons in an area and the number of completed gender affirming surgeries, which can be challenging to track.

In conclusion, this research illuminates the profound impact of anti-transgender legislation on TGD individuals, revealing a complex web of challenges exacerbated by discriminatory laws. Our analysis of crowdfunding data demonstrates how these policies intensify systemic barriers, compelling TGD individuals to seek alternative financial support for basic needs and essential medical care. By documenting the prevalence of campaigns for housing, gender-affirming healthcare, living expenses, lack of insurance coverage, experiences of abuse and mental illness, we provide crucial insights into the real-world effects of these laws. This evidence is vital for informing public discourse, shaping inclusive policies, and guiding healthcare providers and advocacy groups. Ultimately, our study serves as a call to action, emphasizing the urgent need to address policy failures that deny TGD individuals access to essential medical treatments and basic security, contributing to the broader goal of creating more equitable and accessible healthcare systems and social policies for the TGD community.

## Data Availability

All data produced in the present study are available upon reasonable request to the authors
Additionally, all data produced are available online at Gofundme.com.

https://www.gofundme.com/

## Notes

### Competing Interest Statement

The authors have declared no competing interest.

### Funding Statement

This study did not receive any funding.

### Author Declarations

The study used ONLY openly available human data that were originally located at Gofundme.com.

## References

1. Warner DM, Mehta AH. Identifying and Addressing Barriers to Transgender Healthcare: Where We Are and What We Need to Do About It. J Gen Intern Med. 2021;36(11):3559–3561. doi:10.1007/s11606-021-07001-21

2. Attacks on Gender Affirming Care by State Map. Human Rights Campaign. Accessed July 14, 2024. https://www.hrc.org/resources/attacks-on-gender-affirming-care-by-state-map

3. Mapping Attacks on LGBTQ Rights in U.S. State Legislatures in 2024. American Civil Liberties Union. Accessed July 14, 2024. https://www.aclu.org/legislative-attacks-on-lgbtq-rights-2024

4. A Wave of Anti-Transgender Legislation. The New York Times. https://www.nytimes.com/2021/04/20/podcasts/the-daily/transgender-girls-sports-republicans.html. April 20, 2021. Accessed July 14, 2024.

5. Legislation Affecting LGBTQ Rights Across the Country 2021. American Civil Liberties Union. Accessed July 20, 2024. https://www.aclu.org/documents/legislation-affecting-lgbtq-rights-across-country-2021

6. Trans Legislation Tracker: 2022 anti-transgender bills. Accessed July 14, 2024. https://translegislation.com/bills/2022

7. The rise of anti-trans bills in the US. Reuters. https://www.reuters.com/graphics/USA-HEALTHCARE/TRANS-BILLS/zgvorreyapd/. August 19, 2023. Accessed July 30, 2024.

8. Branigin A, Kirkpatrick N. Anti-trans laws are on the rise. Here’s a look at where — and what kind. Washington Post. https://www.washingtonpost.com/lifestyle/2022/10/14/anti-trans-bills/. October 14, 2022. Accessed July 14, 2024.

9. Learn | U.S. anti-trans legislation history. Accessed July 25, 2024. https://translegislation.com/learn

10. Inflection Point. Accessed July 20, 2024. https://trans-legislation.substantial.com/

11. A critical commentary on ‘rapid-onset gender dysphoria’-Florence Ashley, 2020. Accessed July 14, 2024. https://journals.sagepub.com/doi/10.1177/0038026120934693

12. Lockmiller C. Decoding the Misinformation-Legislation Pipeline: an analysis of Florida Medicaid and the current state of transgender healthcare. J Med Libr Assoc JMLA. 2023;111(4):750–761. doi:10.5195/jmla.2023.1724

13. Littman L. Parent reports of adolescents and young adults perceived to show signs of a rapid onset of gender dysphoria. PLOS ONE. 2018;13(8):e0202330. doi:10.1371/journal.pone.0202330

14. Ghorayshi A. Doctors Debate Whether Trans Teens Need Therapy Before Hormones. The New York Times. https://www.nytimes.com/2022/01/13/health/transgender-teens-hormones.html. January 13, 2022. Accessed July 14, 2024.

15. Brandt v. Rutledge, 551 F. Supp. 3d 882 | Casetext Search + Citator. Accessed July 14, 2024. https://casetext.com/case/brandt-v-rutledge-1/

16. Age of Realization and Disclosure of Gender Identity Among Transgender Adults-PubMed. Accessed July 14, 2024. https://pubmed.ncbi.nlm.nih.gov/36935303/

17. Rotondi NK, Bauer GR, Scanlon K, Kaay M, Travers R, Travers A. Nonprescribed Hormone Use and Self-Performed Surgeries: “Do-It-Yourself” Transitions in Transgender Communities in Ontario, Canada. Am J Public Health. 2013;103(10):1830–1836. doi:10.2105/AJPH.2013.301348

18. Willis P, Dobbs C, Evans E, Raithby M, Bishop J. Reluctant educators and self-advocates: Older trans adults’ experiences of health-care services and practitioners in seeking gender-affirming services. Health Expect Int J Public Particip Health Care Health Policy. 2020;23(5):1231–1240. doi:10.1111/hex.13104

19. Clark K, Fletcher JB, Holloway IW, Reback CJ. Structural Inequities and Social Networks Impact Hormone Use and Misuse among Transgender Women in Los Angeles County. Arch Sex Behav. 2018;47(4):953–962. doi:10.1007/s10508-017-1143-x

20. Angraal S, Zachariah AG, Raaisa R, et al. Evaluation of Internet-Based Crowdsourced Fundraising to Cover Health Care Costs in the United States. JAMA Netw Open. 2021;4(1):e2033157. doi:10.1001/jamanetworkopen.2020.33157

21. Barcelos CA, Budge SL. Inequalities in Crowdfunding for Transgender Health Care. Transgender Health. 2019;4(1):81–88. doi:10.1089/trgh.2018.0044

22. Akiki RK, Borrelli MR, Kwan D. Online Crowdfunding Enables Patients’ Access to Gender-Affirming Surgery. Transgender Health. 2021;6(5):240–243. doi:10.1089/trgh.2020.0128

23. Wade M. ‘The giving layer of the internet’: A critical history of GoFundMe’s reputation management, platform governance, and communication strategies in capturing peer-to-peer and charitable giving markets. J Philanthr Mark. 2023;28(4):e1777. doi:10.1002/nvsm.1777

24. GoFundMe | The #1 Crowdfunding and Fundraising Platform. Accessed August 23, 2024. https://www.gofundme.com/

25. Richardson L. beautifulsoup4: Screen-scraping library. Accessed July 25, 2024. https://www.crummy.com/software/BeautifulSoup/bs4/

26. Coding-Qualitative Data Analysis-LibGuides at University of Illinois at Urbana-Champaign. Accessed July 14, 2024. https://guides.library.illinois.edu/qualitative/coding

27. Python Release Python 3.9.0. Python.org. Accessed August 23, 2024. https://www.python.org/downloads/release/python-390/

28. pandas-Python Data Analysis Library. Accessed August 23, 2024. https://pandas.pydata.org/

29. NumPy -. Accessed August 23, 2024. https://numpy.org/

30. Statistical functions (scipy.stats) — SciPy v1.14.1 Manual. Accessed August 23, 2024. https://docs.scipy.org/doc/scipy/reference/stats.html

31. Faletsky A, Han JJ, Lee KJ, et al. Crowdfunding for Gender-Affirming Mastectomy: Balancing Fundraising With Loss of Privacy. Ann Plast Surg. 2022;88(4):372–374. doi:10.1097/SAP.0000000000002953

32. ‘Bye-bye boobies’: normativity, deservingness and medicalisation in transgender medical crowdfunding: Culture, Health & Sexuality: Vol 21 , No 12-Get Access. Accessed July 14, 2024. https://www.tandfonline.com/doi/full/10.1080/13691058.2019.1566971

33. A cross-sectional study of social inequities in medical crowdfunding campaigns in the United States | PLOS ONE. Accessed July 14, 2024. https://journals.plos.org/plosone/article?id=10.1371/journal.pone.0229760

34. US Trans Survey. US Trans Survey. Accessed July 31, 2024. https://ustranssurvey.org/

35. Nakajima K. Bills targeting trans youth are growing more common — and radically reshaping lives. NPR. https://www.npr.org/2022/11/28/1138396067/transgender-youth-bills-trans-sports. November 28, 2022. Accessed July 14, 2024.

36. The Impact of 2023 Legislation on Transgender Youth. Williams Institute. Accessed July 31, 2024. https://williamsinstitute.law.ucla.edu/publications/2023-trans-legislative-summary/

37. Bockting WO, Miner MH, Swinburne Romine RE, Hamilton A, Coleman E. Stigma, Mental Health, and Resilience in an Online Sample of the US Transgender Population. Am J Public Health. 2013;103(5):943–951. doi:10.2105/AJPH.2013.301241

38. Puckett JA, Matsuno E, Dyar C, Mustanski B, Newcomb ME. Mental health and resilience in transgender individuals: What type of support makes a difference? J Fam Psychol. 2019;33(8):954–964. doi:10.1037/fam0000561

39. Breslow AS, Brewster ME, Velez BL, Wong S, Geiger E, Soderstrom B. Resilience and collective action: Exploring buffers against minority stress for transgender individuals. Psychol Sex Orientat Gend Divers. 2015;2(3):253–265. doi:10.1037/sgd0000117

40. Perez-Brumer A, Day JK, Russell ST, Hatzenbuehler ML. Prevalence and Correlates of Suicidal Ideation Among Transgender Youth in California: Findings From a Representative, Population-Based Sample of High School Students. J Am Acad Child Adolesc Psychiatry. 2017;56(9):739–746. doi:10.1016/j.jaac.2017.06.010

41. Nunn R, Parsons J, Shambaugh J. A Dozen Facts about the Economics of the U.S. Health-Care System.

42. Chu J, Nagpal M, Dobberfuhl AD. Utilization and Cost of Gender-affirming Surgery in the United States from 2012-2019. Ann Surg.:10.1097/SLA.0000000000006296. doi:10.1097/SLA.0000000000006296

43. Gender Reassignment Surgery. Accessed July 14, 2024. https://www.bcbst.com/mpmanual/!SSL!/WebHelp/Gender_Reassignment.htm

44. Issue brief: Health insurance coverage for gender-affirming care of transgender patients. Published online 2019.

45. Overview of gender-affirming treatments and procedures | Gender Affirming Health Program. Accessed August 15, 2024. https://transcare.ucsf.edu/guidelines/overview

46. Puckett JA, Cleary P, Rossman K, Newcomb ME, Mustanski B. Barriers to Gender-Affirming Care for Transgender and Gender Nonconforming Individuals. Sex Res Soc Policy J NSRC SR SP. 2018;15(1):48–59. doi:10.1007/s13178-017-0295-8

47. Transgender Workplace Rights. Lambda Legal Legacy. Accessed July 14, 2024. https://legacy.lambdalegal.org/know-your-rights/article/trans-workplace

48. Padula WV, Heru S, Campbell JD. Societal Implications of Health Insurance Coverage for Medically Necessary Services in the U.S. Transgender Population: A Cost-Effectiveness Analysis. J Gen Intern Med. 2016;31(4):394–401. doi:10.1007/s11606-015-3529-6

49. Gonzales G, Henning-Smith C. Barriers to Care Among Transgender and Gender Nonconforming Adults. Milbank Q. 2017;95(4):726–748. doi:10.1111/1468-0009.12297

50. Safer JD, Coleman E, Feldman J, et al. Barriers to healthcare for transgender individuals. Curr Opin Endocrinol Diabetes Obes. 2016;23(2):168. doi:10.1097/MED.0000000000000227

51. White Hughto JM, Reisner SL, Pachankis JE. Transgender stigma and health: A critical review of stigma determinants, mechanisms, and interventions. Soc Sci Med. 2015;147:222–231. doi:10.1016/j.socscimed.2015.11.010

52. Crissman HP, Berger MB, Graham LF, Dalton VK. Transgender Demographics: A Household Probability Sample of US Adults, 2014. Am J Public Health. 2017;107(2):213–215. doi:10.2105/AJPH.2016.303571

53. Gonzales G, Henning-Smith C, Ehrenfeld JM. Changes in health insurance coverage, access to care, and health services utilization by sexual minority status in the United States, 2013-2018. Health Serv Res. 2021;56(2):235–246. doi:10.1111/1475-6773.13567

54. Prohibiting Gender-Affirming Medical Care for Youth. Williams Institute. Accessed July 31, 2024. https://williamsinstitute.law.ucla.edu/publications/bans-trans-youth-health-care/

55. Human Rights Campaign: Florida Boards of Medicine Relentless in Pursuing Anti-Scientific, Harmful, Discriminatory Rule to Deny Care to Transgender Youth. Human Rights Campaign. February 10, 2023. Accessed July 14, 2024. https://www.hrc.org/press-releases/human-rights-campaign-florida-boards-of-medicine-relentless-in-pursuing-anti-scientific-harmful-discriminatory-rule-to-deny-care-to-transgender-youth

56. Alabama SB184 | 2022 | Regular Session. LegiScan. Accessed July 14, 2024. https://legiscan.com/AL/text/SB184/id/2566425

57. Cunningham GB, Watanabe NM, Buzuvis E. Anti-transgender rights legislation and internet searches pertaining to depression and suicide. PLOS ONE. 2022;17(12):e0279420. doi:10.1371/journal.pone.0279420

58. Barbee H, Deal C, Gonzales G. Anti-Transgender Legislation—A Public Health Concern for Transgender Youth. JAMA Pediatr. 2022;176(2):125–126. doi:10.1001/jamapediatrics.2021.4483

59. Turban JL, King D, Kobe J, Reisner SL, Keuroghlian AS. Access to gender-affirming hormones during adolescence and mental health outcomes among transgender adults. PLOS ONE. 2022;17(1):e0261039. doi:10.1371/journal.pone.0261039

60. Wesp LM, Malcoe LH, Elliott A, Poteat T. Intersectionality Research for Transgender Health Justice: A Theory-Driven Conceptual Framework for Structural Analysis of Transgender Health Inequities. Transgender Health. 2019;4(1):287–296. doi:10.1089/trgh.2019.0039

61. Transgender people over four times more likely than cisgender people to be victims of violent crime. Williams Institute. Accessed August 15, 2024. https://williamsinstitute.law.ucla.edu/press/ncvs-trans-press-release/

62. Bowleg L. The Problem With the Phrase Women and Minorities: Intersectionality—an Important Theoretical Framework for Public Health. Am J Public Health. 2012;102(7):1267–1273. doi:10.2105/AJPH.2012.300750

63. Stroumsa D. The State of Transgender Health Care: Policy, Law, and Medical Frameworks. Am J Public Health. 2014;104(3):e31–e38. doi:10.2105/AJPH.2013.301789

64. Trans Experiences Of Homelessness: Disparities, Discrimination, And Solutions – Partnership for Strong Communities. Accessed July 14, 2024. https://pschousing.org/trans-experiences-of-homelessness-disparities-discrimination-and-solutions/

65. Rich AJ, Scheim AI, Koehoorn M, Poteat T. Non-HIV chronic disease burden among transgender populations globally: A systematic review and narrative synthesis. Prev Med Rep. 2020;20:101259. doi:10.1016/j.pmedr.2020.101259

66. Cooper K, Mandy W, Butler C, Russell A. The lived experience of gender dysphoria in autistic adults: An interpretative phenomenological analysis. Autism. 2022;26(4):963–974. doi:10.1177/13623613211039113

67. Baker K, Restar A. Utilization and costs of gender-affirming care in a commercially insured transgender population. Journal of Law, Medicine & Ethics. 2022;50(3):456–470. doi:10.1017/jme.2022.87

68. Olansky E, Lee K, Handanagic S, Trujillo L. Nonprescription hormone use among transgender women-national HIV behavioral surveillance among transgender women, seven urban areas, United States, 2019–2020. Centers for Disease Control and Prevention. January 25, 2024. Accessed September 1, 2024. https://www.cdc.gov/mmwr/volumes/73/su/su7301a4.htm.

69. Rotondi NK, Bauer GR, Scanlon K, Kaay M, Travers R, Travers A. Nonprescribed hormone use and self-performed surgeries: “do-it-yourself” transitions in transgender communities in Ontario, Canada. American Journal of Public Health. 2013;103(10):1830–1836. doi:10.2105/ajph.2013.301348

